# Spatial Correlation of Particulate Matter Pollution and Death Rate of COVID-19

**DOI:** 10.1101/2020.04.07.20052142

**Authors:** Ye Yao, Jinhua Pan, Weidong Wang, Zhixi Liu, Haidong Kan, Xia Meng, Weibing Wang

## Abstract

The Coronavirus (COVID-19) epidemic, which was first reported in December 2019 in Wuhan, China, has caused 3,314 death as of March 31, 2020 in China. This study aimed to investigate the spatial associations of daily particulate matter (PM) concentrations with death rate of COVID-19 in China. We conducted a cross-sectional analysis to examine the spatial associations of daily PM2.5 and PM10 concentrations with death rate of COVID-19 in China through multiple linear regression method. We found that COVID-19 held higher death rates with increasing concentration of PM2.5 and PM10 levels in the spatial scale, which may affect the process of patients developed from mild to severe and finally influence the prognosis of COVID-19 patients.

## Introduction

SARS-CoV-2 is a new emerge of coronavirus that poses immense challenges to global health and the economy. As of April 3, 2020, a total of 1,042,588 COVID-19 cases have been confirmed in 199 countries worldwide. In China, a total of 82,772 confirmed cases and 3,327 deaths were recorded. World Health Origination (WHO) announced on March 11 that the assessment of the COVID-19 outbreak has been a global pandemic.

The hypothesis that air pollution can act both as a vector of the infection and as a worsening factor of the health impact of the pandemic in progress has been raised recently. Short-term exposure to ambient particulate matter (PM) pollution has been associated with increasing risks of mortality and morbidity of cardiopulmonary diseases worldwide^1,2^, and the adverse effects are significantly higher for people who are older or with underlying medical conditions^3^, among which the fatality of COVID-19 is also higher. While previous studies suggested that short-term exposure to PM could aggravate respiratory symptoms and increase the emergency visits of patients with asthma and chronic obstructive pulmonary disease (COPD) ^4^, the European Public Health Alliance have recently reported that air pollution is likely to increase the death rate of SARS-CoV-2 in cities^5^. Air pollution, especially PM pollution, was approved to be also positively associated case fatality of severe acute respiratory syndrome (SARS) in China^6^.

However, few studies have estimated and quantified the short-term effects of air pollutants on early deaths from COVID-19. While the extent of correlation is less known, this study aimed to investigate the association between death rate of COVID-19 and PM_2.5_ and PM_10_ pollutions in a spatial scale.

## Methods

We conducted a cross-sectional analysis to examine the spatial associations of daily PM_2.5_ and PM_10_ concentrations with death rate of COVID-19 in China through multiple linear regression method. The COVID-19 confirmed cases and deaths information in China was collected from the National Health Commission and the Provincial Health Commissions of China. We calculated death rate, which was defined as cumulative death counts divided by cumulative confirmed cases, for 49 cities including Wuhan, other 15 cities inside Hubei and 33 cities outside Hubei, with no less than 100 cases as of March 22 (few confirmed cases and deaths were reported afterwards). Daily PM_2.5_ and PM_10_ data were collected from the National Urban Air Quality Publishing Platform (http://106.37.208.233:20035/), which is administered by China’s Ministry of Environmental Protection. And meteorological data including daily mean temperature and relative humidity were collected from the China Meteorological Data Sharing Service System. Gross Domestic Product (GDP) per capita, hospital beds and population size were obtained from statistical yearbook of each province.

## Results

As of March 22, the mean death rate of COVID-19 was 1.7% among the 49 cities in China, with a range of 0.0%-5.0%. And the mean ± standard deviation and range were (51.2±20.9, 20.8-96.9 μg/m^3^ & 62.1±22.6, 30.8-115.1 μg/m^3^) for PM_2.5_ & PM_10._ There were no significant PM_2.5_ or PM_10_ differences between cities outside and inside Hubei (t=0.48, *p*=0.63 & t=0.63, *p*=0.53). Death rate of cities inside Hubei except Wuhan was significantly lower than that of Wuhan (t=6.70, *p*=5.0×10^−5^), while death rate of cities outside Hubei was even lower than those inside Hubei (t=8.26, *p*=2.6×10^−10^).

After adjustment for temperature, relative humidity, GDP per capita and hospital beds per capita, COVID-19 death rate was positively associated with concentration of PM_2.5_ (**Figure 1A**, *χ*^2^ =13.10, p=0.011) and PM_10_ (**Figure 1B**, χ^*2*^ =12.38, p=0.015), suggesting that COVID-19 held higher death rates with increasing concentration of PM_2.5_ and PM_10_ levels in the spatial scale. In addition, we did not find significance in the association between GDP per capita, hospital beds per capita, temperature or relative humanity and COVID-19 death rate (χ^2^=6.02, *p*=0.20 & χ^*2*^ =3.68, *p*=0.45 & χ^*2*^ =3.76, *p*=0.44 & χ^*2*^ =7.21, *p*=0.13).

**Figure 1.**
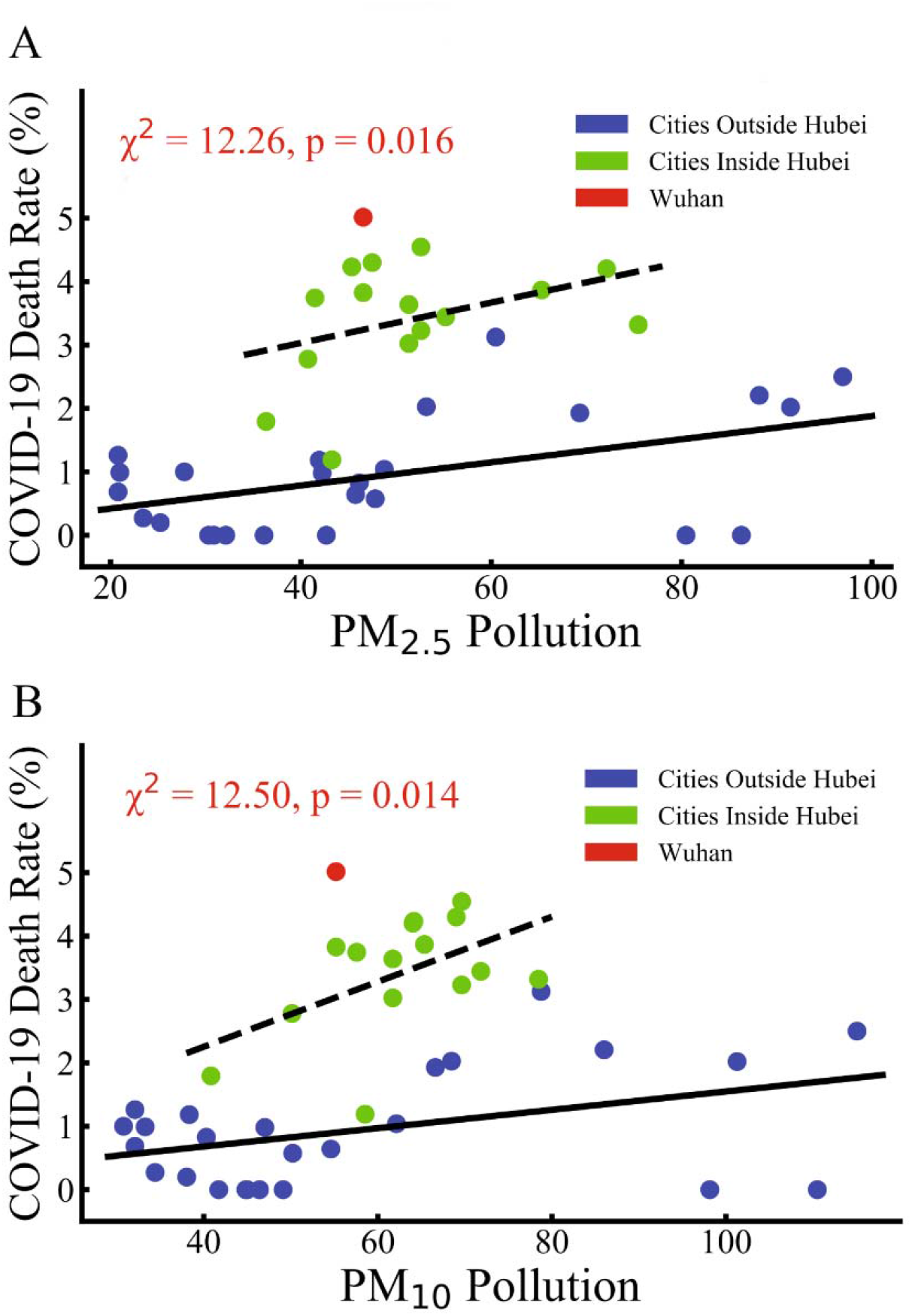
Death Rate Versus PM_2.5_ & PM_10_ pollution. **Figure 1A:** Death Rate was positively associated (Meta *χ*^2^=13.10, p=0.011) with PM_2.5_ in cities outside Hubei (blue points, r=0.56, p=0.005) and those inside Hubei except Wuhan (green points, r=0.33, p=0.26) pollution. GDP per capita and hospital beds per capita effects were removed during statistical analysis. Figure 1B: Death Rate was positively associated (Meta χ ^2^=12.38, p10 =0.015) with PM_10_ in cities outside Hubei (blue points, r=0.48, p=0.019) and those inside Hubei except Wuhan (green points, r=0.47, p=0.11) pollution. GDP per capita and hospital beds per capita effects were removed during statistical analysis.

## Discussions

Our results demonstrate that the death rate of coronavirus disease (COVID-19) has a strong association with PM_2.5_ and PM_10_, whether in Hubei province or other cities in China. Our results are consistent with previous studies of SARS^6^. COVID-19 is caused by SARS-CoV-2, which shares 79.6% sequence identity to SARS-CoV and has the same cell entry receptor - angiotensin converting enzyme II - as SARS-CoV. Limited studies have researched on the associations between air pollution and SARS which only lasted for half year in 2003 and only a few Chinese cities had adequate death data for analysis. A previous one reported that moderate air pollution index (APIs), which was dominated by PM_10_ in China in 2003, had an 84% increased risk of dying from SARS compared to those from regions with low APIs, and SARS patients from regions with high APIs were twice as likely to die from SARS compared to those from regions with low APIs. Other previous studies demonstrated that long-term or short-term exposure to PM_10_ and PM_2.5_ might damage the lung functions^7-9^. The mechanism of the effects from PM exposure on respiratory outcomes might be caused by activation of inflammatory pathways in the small airways in response to PMs, leading to the recruitment of inflammatory cells^10^. These biological mechanisms might potentially influence the prognosis of COVID-19 patients.

Considering the fact that the patients died from COVID-19 are likely to be critically ill, most of them might stay in Intensive Care Unit (ICU) for treatment. We speculate that the effects of PM_2.5_ and PM_10_ on death mainly affect the progress of patients from mild to severe and prognosis. There is a need to increase our efforts in the control of air pollutant emissions, especially regarding the situation that the COVID-19 epidemics may come back at any time.

The study was limited to a short period of season with less variation of air pollution. However, the correlation of trends of death and air pollution is quite convincing. Given the ecological nature of study, other city-level factors, such as implementation ability of COVID-19 control policy, urbanization rate, and availability of medical resources, may affect the death rate of COVID-19 and confound our findings, however, we control the GDP per capita and hospital beds per capita increased the credibility of the results.

## Data Availability

Daily PM2.5 and PM10 data were collected from the first link below.
COVID-19 confirmed cases and deaths information be obtained from the second and third links.

http://106.37.208.233:20035/

http://www.nhc.gov.cn/xcs/xxgzbd/gzbd_index.shtml

http://wjw.hubei.gov.cn/bmdt/ztzl/fkxxgzbdgrfyyq/

## Author contributions

Dr. Ye Yao, Jinhua Pan, Weidong Wang and Zhixi Liu contributed equally. Dr. Weibing Wang and Dr. Xia Meng contributed equally.

Concept and design: Ye Yao, Xia Meng, and Weibing Wang.

Acquisition, analysis, or interpretation of data: Ye Yao, Jinhua Pan, Zhixi Liu,

Haidong Kan and Weidong Wang.

Drafting of the manuscript: Ye Yao, Jinhua Pan, Zhixi Liu.

Critical revision of the manuscript for important intellectual content: Haidong Kan,

Xia Meng and Weibing Wang.

Statistical analysis: Ye Yao and Xia Meng.

## Acknowledgements

This study was sponsored by the Bill & Melinda Gates Foundation (Grant No. OPP1216424).

